# Perioperative oxygen concentration and postoperative organ damage in pediatric neurosurgery: a retrospective cohort study

**DOI:** 10.1101/2024.11.12.24317154

**Authors:** Zhengfang Hu, Chenwei Wang, Huiwen Wang, Kangda Zhang

## Abstract

**Purpose:** To investigate the correlation between perioperative oxygen concentration and postoperative organ damage and other adverse reactions in children undergoing neurosurgery.

**Materials and methods:** This study was a single-center retrospective cohort study that included 512 children undergoing neurosurgery, aged ≤14 years old, and with an operative duration ≥2 hours who were transferred to the Postanesthesia Care Unit (PACU) of the Department of Anesthesiology, Beijing Tiantan Hospital, postoperatively during the period of October 2018 to September 2023. The medical records of the included children during hospitalization were meticulously reviewed and systematically collected through the Hospital Information System (His).The area under oxygen concentration-time curve AUC_FIO2_= the product of intraoperative oxygen concentration (percentage) and duration of anesthesia (minutes) was calculated, and then 512 children were divided into 3 groups according to the trichotomies of the AUC_FIO2_ (Group1:AUC_FIO2_<8720%min, Group2: 8720% min ≤ AUC_FIO2_ ≤ 13800% min, Group 3: AUC_FIO2_ > 13800% min) for inclusion in the data analysis.

**Results:** The incidence of Emergency delirium was 24%, 26.9% and 19.4%, and the incidence of acute kidney injury was 0.6%, 2.3% and 0.6% in the three groups, respectively. There was no significant difference between the three groups (P> 0.05). The incidence of postoperative pulmonary complications in the three groups was 20.6%, 20.8% and 31.5%, respectively, and there was a statistically significant difference between the three comparisons (P<0.05). There were no significant differences in nausea, vomiting, hypoxemia, VAS score and PACU residence time among the three groups during PACU (P>0.05), and there was a statistically significant differences between infections and hospital stay during hospitalization (P<0.05).

**Conclusions:** Although the oxygen concentration AUC_FIO2_ was not correlated with the occurrence of both postoperative AKI and ED, the longer the exposure time of conventional oxygen administration strategy, the higher the incidence of postoperative pulmonary complications and postoperative infections, and the longer the hospitalization time of children.

## Introduction

Almost all patients undergoing general anesthesia require routine intraoperative oxygen administration to counteract the diminished gas exchange induced by the various intraoperative anesthetics and analgesics and to ensure adequate oxygenation from the time of anesthesia intubation to the time of awakening from anesthesia. Children, however, are more likely to use high concentration of oxygen undergoing surgical treatment due to their smaller functional residual gas volumes and higher oxygen consumption ^[1]^. However, oxygen is a double-edged sword, and in the 1990s, adverse outcomes of hyperoxygenation (supraphysiologic oxygen delivery) in critically ill neonates were first described^[2,3]^, culminating in national guidelines recommending the administration of room-air-concentrated oxygen for critically ill neonates^[4]^. These findings prompted further research to evaluate the potential adverse effects of hyperoxia on other populations and different organs. 2022, McIlroy’s team ^[5]^, found in a large, heterogeneous, and contemporary cohort of adult patients undergoing a wide range of surgical procedures requiring general anesthesia, the incidence of postoperative AKI, myocardial injury, and lung injury was each higher in patients exposed to increased supraphysiological oxygen administration (defined as the area under the curve of the fraction of inspired oxygen above air during minutes when the hemoglobin oxygen saturation was greater than 92%)during surgery, indicating that the intensity and duration of exposure to supraphysiological oxygen delivery affects oxygen toxicity.

Given that children have distinct physiological responses compared to adults, which may alter their susceptibility to hypoxia or hyperoxia during surgical procedures. however, the existing literature primarily focuses on adult populations and the studies exploring the relationship between perioperative inspired oxygen concentration and the prognosis of children’s surgeries are limited, leaving a significant gap in our understanding that the influence of intraoperative oxygen exposure time and intensity on various organs of pediatric patients.

The pediatric neurosurgical operations are characterized by long operation duration and prolonged anesthesia times, which can lead to varying levels of oxygenation throughout the procedure, so we propose to select children undergoing neurosurgical procedures as the research subjects and retrospectively observe the effects of the current clinical routine oxygen supply strategies on postoperative cardiac, pulmonary, and renal injuries and incidences of Emergency delirium during the awakening period in children. By systematically collecting data from medical records regarding intraoperative oxygen concentrations and durations alongside patient recovery metrics post-surgery—such as vital signs monitoring and organ function tests—we hope to elucidate potential correlations. Ultimately, enhancing our understanding in this area may contribute significantly toward improving overall patient safety and outcomes in pediatric neurosurgery.

## Methods

### Setting, patient population, and study design

This is a retrospective observational cohort study conducted from October 2018 to September 2023 approved by the ethics committee of the Beijing Tiantan Hospital (KY-2024-037 ), Capital Medical University. The children who had neurosurgery and were sent to the PACU of the Anesthesia Department of Beijing Tiantan Hospital for after-surgery care were retrieved and analyzed. The 512 children data included in the study were based on the following inclusion criteria of: ➀ age 14 or younger; ➁ general anesthesia; ➂ the types of surgeries were intracranial tumor removal, cerebrovascular malformation removal and spinal cord and spinal column tumor removal; ➃ the surgery lasted for 2 hours or longer. The exclusion criteria were: ➀ missing data in the medical records; ➁ SPO_2_ was less than 90% for more than 3 minutes during the operation; ➂ having another surgery within 90 days.

### Clinical protocols

#### Patient and public involvement

This was a de-identified chart review and as such no patients were directly involved.

#### Data collection

The medical records of the included children during hospitalization were meticulously reviewed and systematically collected through the Hospital Information System (His). The baseline data encompassed a variety of demographic and clinical parameters, including age, gender, ASA classification, body mass, anesthetic medication usage patterns, duration of anesthesia procedures, intraoperative oxygen inhalation concentration levels, postoperative diagnosis details, and surgical approach utilized. Several vital signs and recovery metrics were documented including blood pressure, heart rate, oxygen saturation levels monitored throughout the procedure and recovery phase, pain scores assessed using standardized scales at various intervals post-surgery.

Furthermore, specific adverse events such as infections—both wound infections confirmed by bacterial culture results obtained from samples taken around the surgical site—and intracranial infections diagnosed through lumbar puncture alongside cerebrospinal fluid examination findings—were carefully recorded. Postoperative pulmonary complications were identified based on diagnostic imaging reports from chest X-rays or chest CT after surgery or changes in lung markings compared to preoperative findings within 72 hours ^[8]^.Changes in renal function were evaluated by calculating the difference between the first postoperative serum creatinine value and the last preoperative serum creatinine value, Postoperative AKI was defined as the difference between the first serum creatinine value and the last preoperative serum creatinine value≥0.3mg/dl within 72h after surgery[7].

Additionally, attention was given to monitoring for emergence delirium—a condition characterized by confusion or agitation upon waking from anesthesia—which was specifically tracked within the Post-Anesthesia Care Unit (PACU) throughout each child’s postoperative hospital stay. To assess emergence delirium quantitatively among pediatric patients in PACU settings effectively, healthcare professionals employed the Pediatric Anesthesia Emergence Delirium (PAED) scale. This validated tool evaluates behaviors exhibited by children after surgery across five distinct categories: awareness level upon awakening from anesthesia; ability to engage with caregivers; emotional state; motor activity level; and overall behavior quality postoperatively. Each behavior was rated on a five-level scale ranging from 0 (indicating no symptoms) to 4 (indicating severe symptoms). The total score derived from these evaluations could reach a maximum of 20 points; an ED diagnosis would be established when a child’s total PAED score met or exceeded 12 points ^[6]^.

Based on McIlroy’s study, the area under the oxygen concentration-time curve (AUC_FIO2_) was first calculated as the product of the intraoperative oxygen concentration (expressed in percentage) and the duration of anesthesia (measured in minutes). And then according to the tertiles of AUC_FIO2_ values, 512 children were classified into 3 groups: Group 1 consisted of those with an AUC_FIO2_ value < 8720% min, Group 2 included participants whose 8720% min ≤ AUC_FIO2_ ≤ 13800% min, Group 3 encompassed individuals with an AUC_FIO2_≥ 13800% min for data analysis.

#### Definitions

The primary outcome indicators were the incidence of ED in PACU, the incidence of postoperative Acute Kidney Injury (AKI), and the incidence of postoperative pulmonary complications in the three groups. Secondary outcome indicators were nausea, vomiting, Visual Analogue Scale(VAS) pain score and hypoxemia in the PACU, infection and length of stay(LOS)during the hospitalization. Hypoxemia during PACU was defined as SpO2 < 90%.

#### Statistical methods

Multiple interpolation was performed for variables with missing proportions of less than 5%, and all continuous variables were first tested for normality, with non-normally distributed measurements expressed as medians (quartiles) and counts expressed as cases (percentage). Kruskal-Wallis rank sum test was used for intergroup comparisons of quantitative data, and chi-square test was used for categorical data. In addition, to further explore the relationship between AUCFiO2 and outcomes, variables with P < 0.1 in univariate analysis were included in the multivariate Logistic regression model. A two-sided P < 0.05 was considered statistically significant, and all statistical analyses were completed on R 4.3.3 software.

## Results

### Baseline characteristics and surgical factors in three groups of children

The baseline characteristics and surgical of the 512 children are shown in Table 1.There were no statistically significant differences in baseline data such as gender (P = 0.484)and intraoperative oxygen saturation among(P =0.249) the three groups, while there were statistically significant differences in age, body mass index, duration of surgery, type of surgery, intraoperative blood loss, colloid infusion, erythrocyte transfusion, plasma transfusion, preoperative Hb and preoperative serum creatinine value among the three groups (P =0.001),which was considered to be related to the subgroups.

**Table 1.**
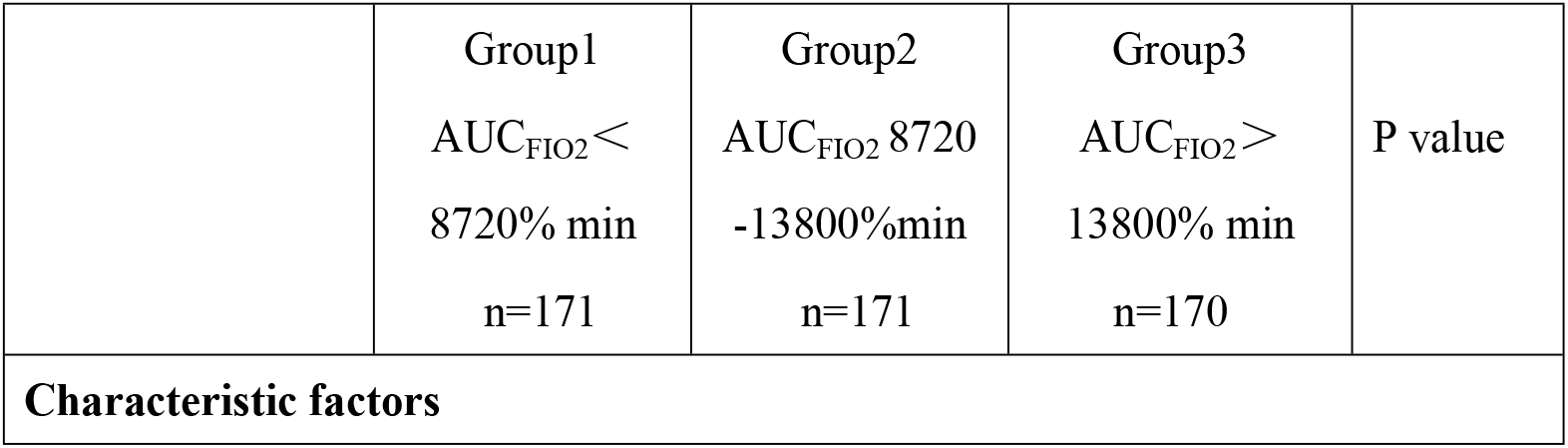
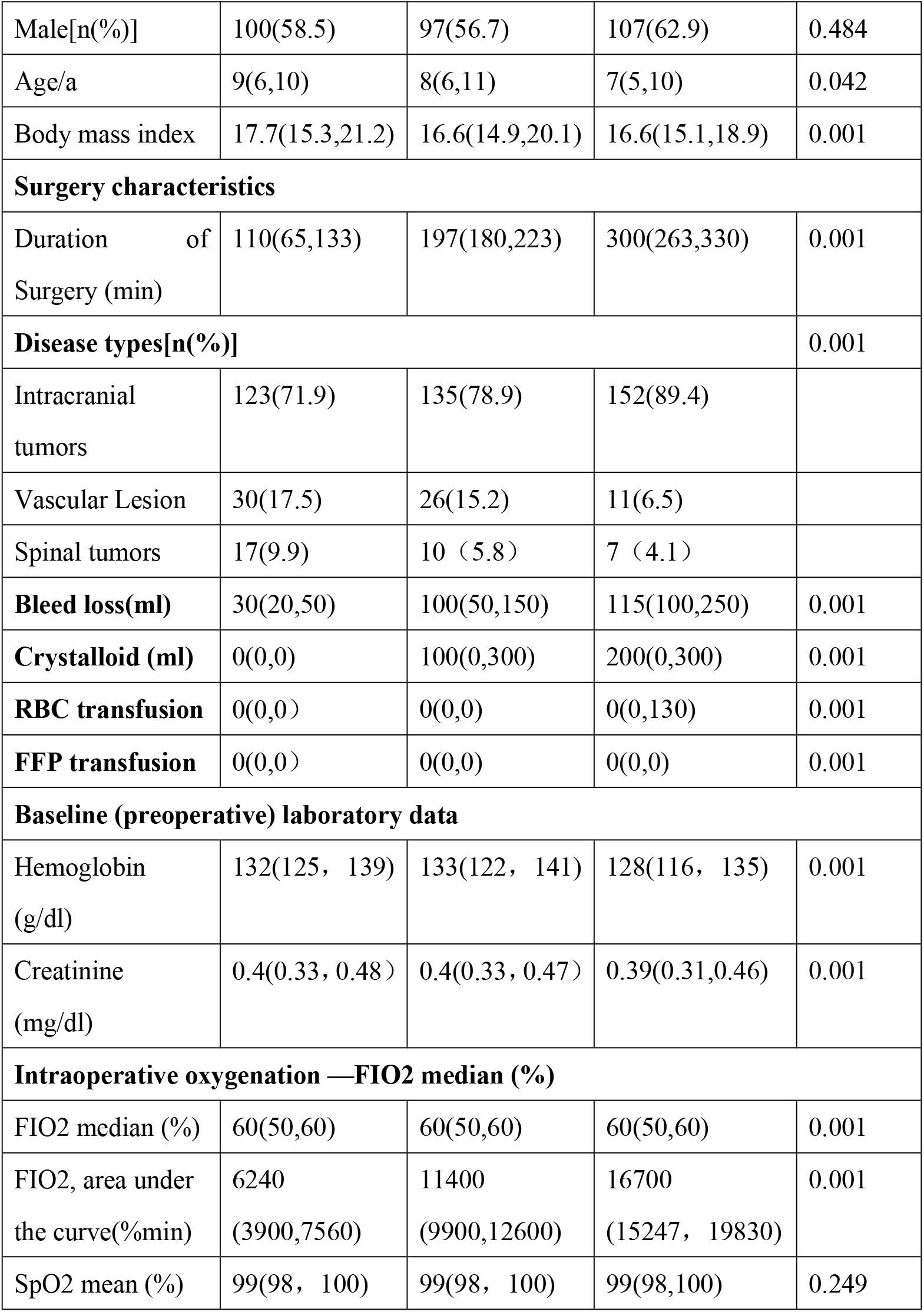
:Comparison of baseline characteristics and surgical factors between the three groups [M(P25,P75)]

### Comparison of primary and secondary outcomes among the three groups

The primary and secondary outcomes as shown in Table 2.The incidence of ED in PACU was 24%, 26.9%, and 19.4% respectively and there was no significant difference among the three groups (p=0.258). The incidence of AKI was 0.6%, 2.3%, and 0.6%, respectively and also there was no significant difference among the three groups (p=0.221). The incidence of postoperative pulmonary complications was 20.6%, 20.8%, and 31.5% in the three groups, respectively, and there was a statistically significant difference between the three comparisons(P=0.028). There was no statistical difference in the incidence of nausea(P=0.579), vomiting(P=0.749), hypoxemia(P=0.606), VAS score(P=0.553), and Resident time in PACU(P=0.195) among the three groups, but there was a statistically significant difference in the comparison of infections(P=0.001), extubation time (P=0.012)and hospital stay during hospitalization(P=0.001).

**Table 2.**
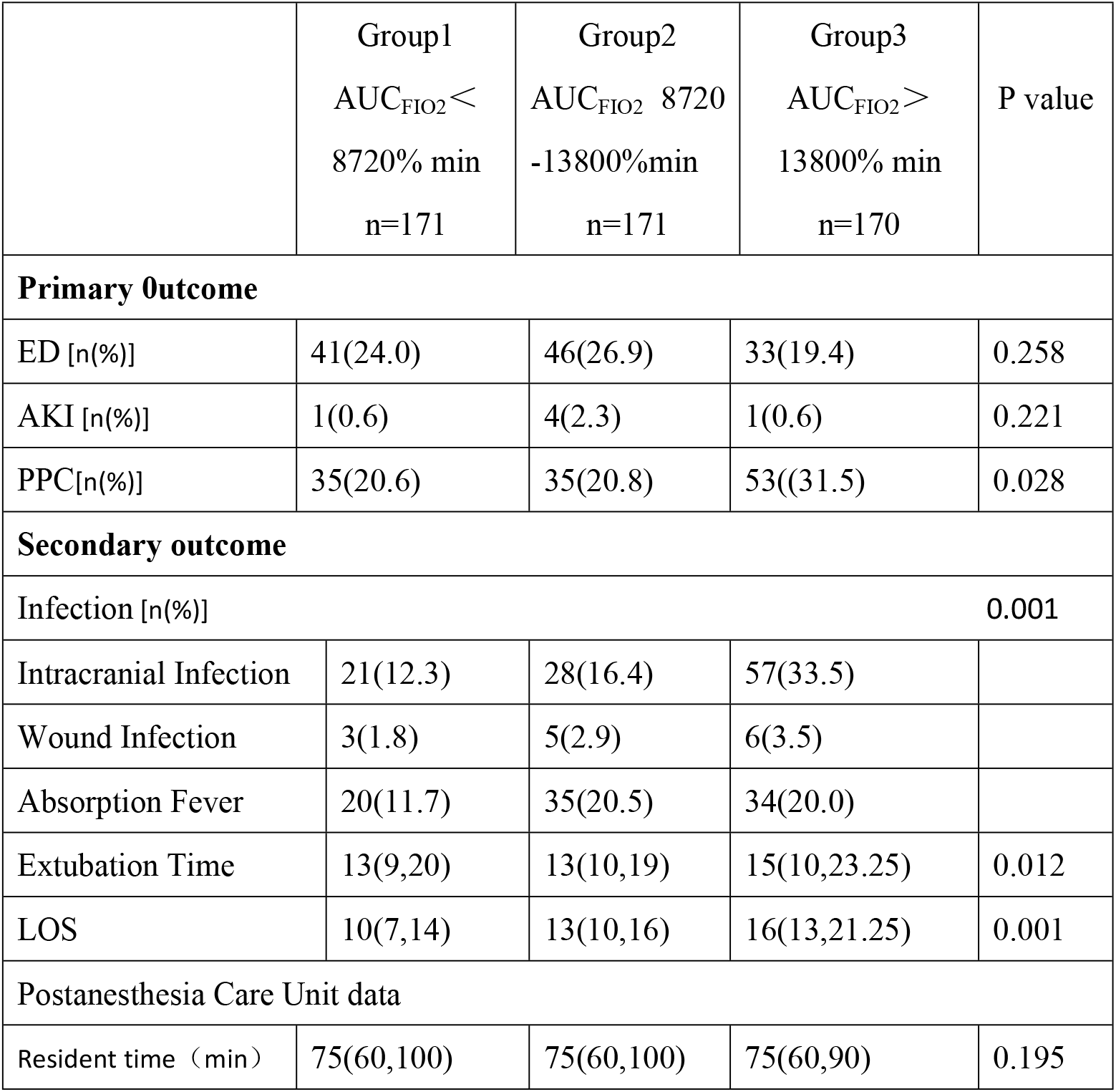
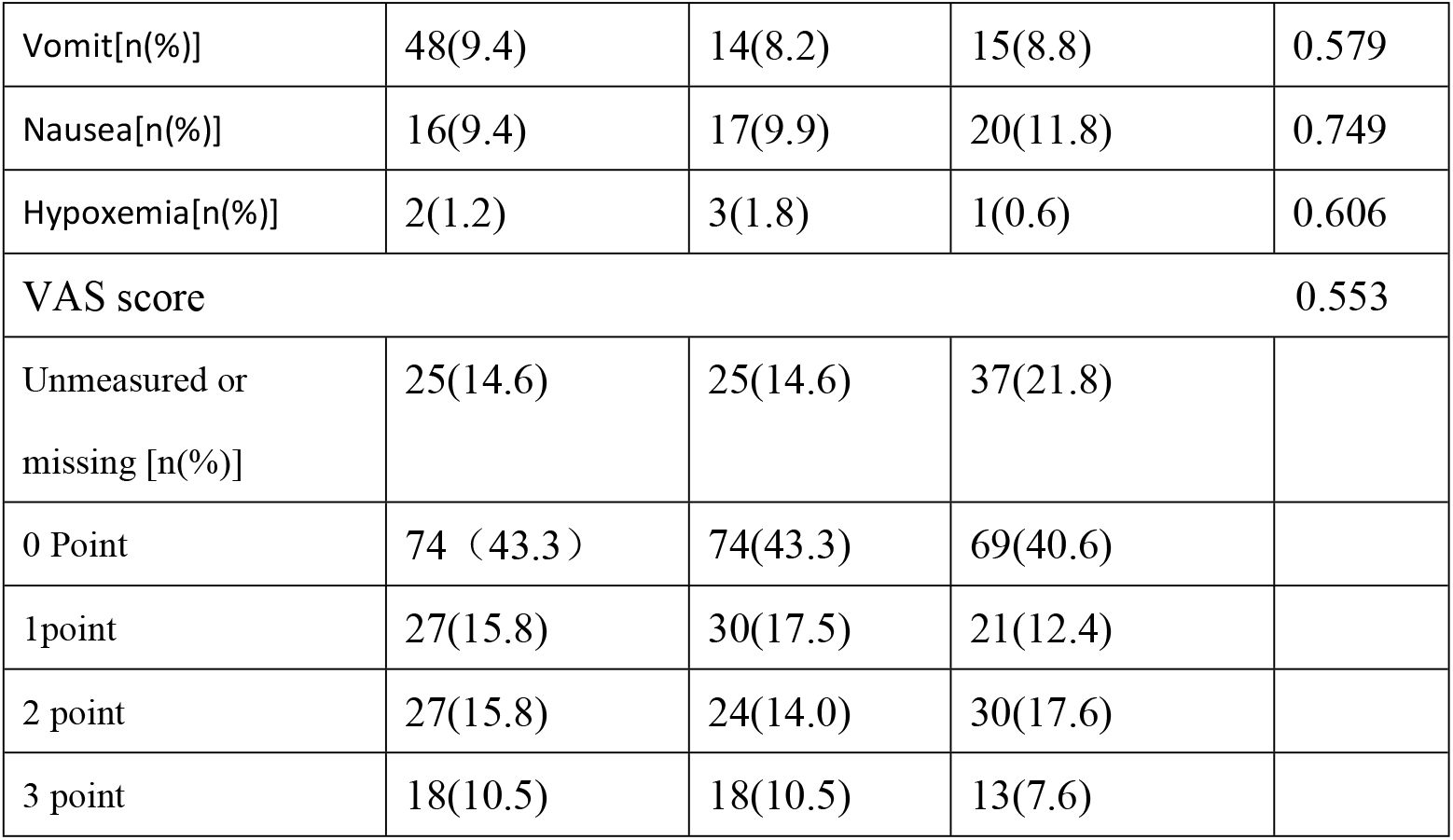
: Comparison of Primary and Secondary outcome between the three groups.

### Correlation analysis between AUC_FIO2_ and primary outcome

Univariate Logistic regression showed that AUC_FIO2_ was negatively correlated with the development of postoperative AKI, but not correlated with ED. However, after adjusting for variables of baseline imbalance, multivariate Logistic regression showed that AUC_FIO2_ was not associated with the occurrence of postoperative AKI or ED (as shown Table 3).

**Table 3.**
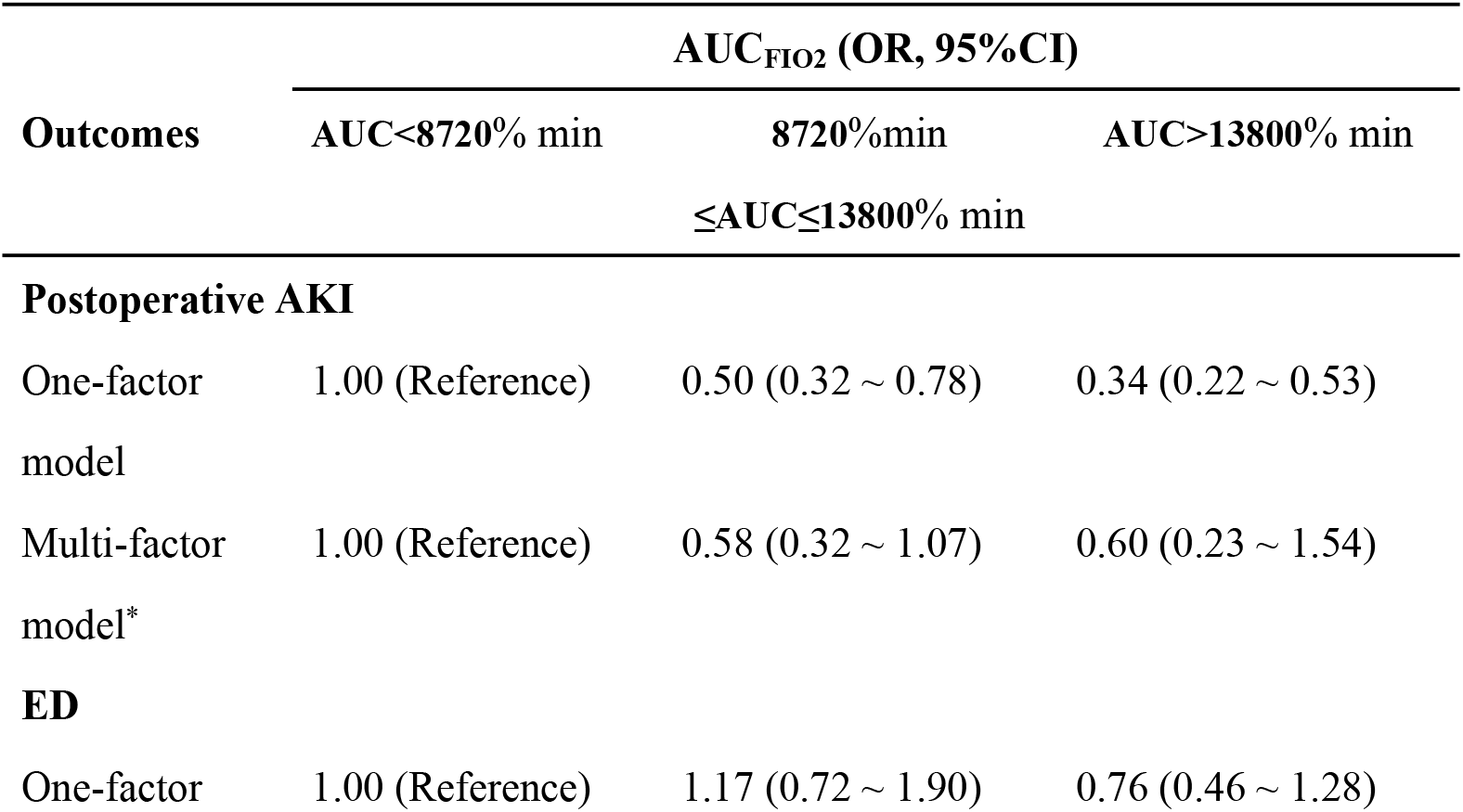
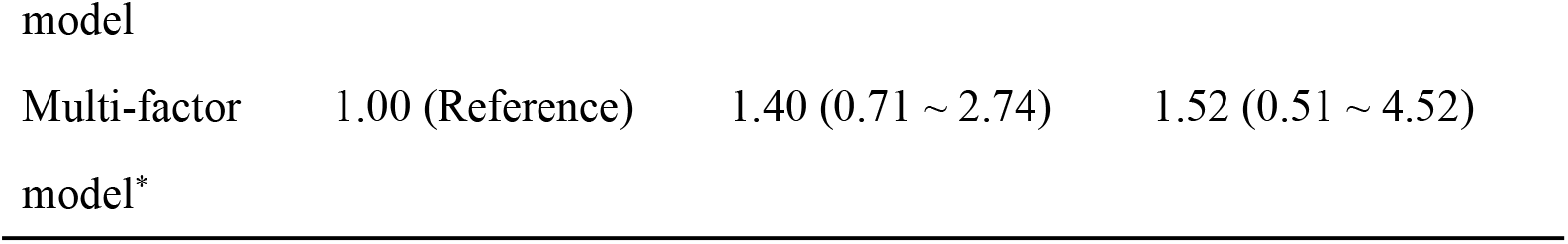
Correlation analysis of AUC_FIO2_ and postoperative AKI and ED * Correcting factors:Postoperative infection、Operation site、Duration of Surgery、Extubation time、Hemoglobin、Body mass index、Age、Blood loss、Crystalloid、RBC transfusion、FFP transfusion # Correcting factors : Postoperative infection, Operation site, Duration of Surgery, Extubation time, Hemoglobin, Body mass index, Age, Blood loss, Crystalloid、RBC transfusion、FFP transfusion、AKI

## Discussion

This study is a retrospective cohort investigation aimed at exploring the influences of intraoperative oxygen exposure duration and intensity on the postoperative conditions of organs such as the brain, lungs, and kidneys in children undergoing neurosurgical procedures. The research results revealed that although the oxygen concentration AUC_FIO2_ had no correlation with the occurrence of postoperative AKI and ED, the longer the exposure time of the conventional oxygen supply strategy, the higher the incidence of postoperative pulmonary complications and postoperative infections in children, and the significantly longer the hospital stay.

Exposure to oxygen concentrations above the normal level might lead to an increase in the production of reactive oxygen metabolites such as hydrogen peroxide, nitric oxide, superoxide, and reactive hydroxyl groups, disrupting the antioxidant balance and thereby promoting lipid peroxidation, impaired protein synthesis, inactivation of enzymes and nucleic acids, DNA mutations, disrupted mitochondrial calcium homeostasis, and increased membrane permeability. It has been regarded as one of the causes of lung, myocardial, and kidney injuries ^[9-11]^. The VISION study suggested that for every 10% increase in intraoperative oxygen inhalation concentration, the risk of postoperative cardiac injury increased by 1.17 times ^[12]^. A retrospective study reported that intraoperative hyperoxia was closely related to acute kidney injury after cardiac surgery^[13]^. Multiple studies have reported that intraoperative hyperoxic cerebral reperfusion injury significantly increased the incidence of early postoperative POCD and postoperative delirium ^[14,15]^. Due to the underdeveloped ROS scavenging mechanism, neonates and children seem to be more susceptible to the multisystem effects of hyperoxia ^[16-18]^. The existing literature observing the relationship between hyperoxemia in children and prognosis is limited and mainly observational. In a Meta-analysis evaluating the relationship between hyperoxemia and mortality in critically ill children, among the 12 studies included by the authors, 7 studies reported that hyperoxemia was closely related to an increased mortality rate in children, while the other 5 studies did not find this association, but their sample sizes were relatively small. Several studies found that this association still persisted after adjusting for demographic characteristics and disease severity, further confirming the potential harmful effects of hyperoxemia. However, the exact threshold at which hyperoxemia becomes harmful remains unclear. Therefore, the authors recommend that clinicians should be rational and cautious when using oxygen therapy for critically ill children ^[19]^. In a prospective observational study by Ayten et al., children undergoing corrective surgery for coarctation of the aorta were used as the observation subjects to compare the differences in postoperative related complications, ICU stay time, and hospital stay time between children who received 80% and 60% oxygen during anesthesia induction. The results showed that although no related complications were recorded in both groups of children after surgery, the ICU stay and hospitalization time of children in the hyperoxia group were significantly longer than those in the hypoxia group ^[20]^.

The results of our retrospective study indicated that the median FiO2 during surgery in 512 children was 60% [50%, 60%], and the median AUC_FiO2_ was 11,400% min [7,515% min, 15,247% min]; the median FiO_2_ during surgery in the David team was 54% [47.5%, 60%], and the median AUC_FiO2_ was 7,951% min [5,870% min, 11,107% min]. Compared with the David team, the median AUC_FiO2_ in our study was significantly larger, which was related to our selection of children undergoing neurosurgical procedures as the observation subjects. The surgery time was long, so the intensity of supra-physiological oxygen exposure (AUC_FiO2_) was high. Different from the research results of the David team, we did not observe statistically significant differences in the incidences of postoperative acute kidney injury and postoperative ED among the three groups of children, which might be related to the small functional residual capacity and high oxygen consumption in children ^[17]^. However, the three groups of children had high incidences of postoperative pulmonary complications and postoperative infections, prolonged hospital stay, and we believe that the reasons for this are partly related to the condition of the children and the other possible reason may be consistent with the assumption of the David team, that exposure intensity and duration affect oxygen toxicity, assuming that exposure intensity and duration are biologically interchangeable, and there is a connection between excess physiological oxygen exposure duration and organ damage. High fractional inspired oxygen, especially ≥ 60%, leads to excessive production of reactive oxygen species in alveolar mitochondria, thereby destroying the normal clearance mechanism and causing cell death ^[21,22]^.

This study has certain limitations: Firstly, this is a single-center retrospective study with inherent limitations such as case selection bias. Additionally, only 1% of the included children had troponin testing after surgery. Therefore, this indicator was excluded and not subjected to statistical analysis. With a larger sample size, perhaps positive findings could have been obtained. Thirdly, postoperative brain tissue resection or injury might affect the early mental state after neurosurgery. Given the ethical limitations of conducting prospective randomized controlled clinical studies in pediatric patients, it is unlikely that the question of the appropriate oxygen concentration during surgery for children undergoing surgery can be directly answered in a prospective randomized controlled study. Therefore, the conclusions of this study may have certain reference value for the intraoperative respiratory management of children undergoing long surgeries. Compared with the conventional oxygen supply strategy, large-scale trials evaluating conservative oxygen supply strategies and targets are more likely to be supported.

## Conclusion

We found that although the oxygen concentration AUC_FIO2_ was not correlated with the occurrence of both postoperative AKI and ED, but the longer the exposure time of conventional oxygen administration strategy, the higher the incidence of postoperative pulmonary complications and postoperative infections, and the longer the hospitalization time of children. This suggests that more stringent oxygen administration strategy and respiratory management measures should be adopted during prolonged pediatric surgery to reduce respiratory related complications and length of hospital stay.

## Data Availability

All relevant data are within the manuscript and its Supporting Information files

## Supporting information

